# Two-year interview and match outcomes of otolaryngology preference signaling

**DOI:** 10.1101/2022.07.12.22277302

**Authors:** C.W. David Chang, Marc C. Thorne, Sonya Malekzadeh, Steven D. Pletcher

## Abstract

**Objective:** To present the first year-over-year data comparison of preference signaling for residency interviews in the otolaryngology application marketplace.

**Study Design:** Cross-sectional study conducted over 2 application cycles

**Setting:** Otolaryngology training programs in the United States.

**Methods:** Otolaryngology residency applicants were invited to participate in preference signaling during the 2021 and 2022 application cycles. Submissions were collected using a web-based interface. The distribution of signals among programs was evaluated descriptively and in relationship to program rankings. Surveys were sent to applicants to assess general attitudes and the number of interview invitations received from signaled and non-signaled programs. Surveys were sent to programs to evaluate use of signals and the impact on Match results.

**Results:** Programs received a range of signals from 0-66, with 50% of signals going to 24% of programs, which was similarly found in 2021. Programs of higher rank tended to receive more signals. Overall, greater than 87% of surveyed applicants received an interview offer from at least one program they signaled. In 2021 and 2022, applicants were 2.6 times more likely to get an interview from a signaled program than from a comparator non-signaled program. A greater positive impact on interview offer rate was seen for less competitive applicants. Signaling was viewed favorably by the vast majority surveyed applicants and programs.

**Conclusions:** Preference signaling for otolaryngology residency interviews demonstrates a promising mechanism to improve applicant visibility to programs during the application cycle. This impact is consistent over 2 application cycles.

## INTRODUCTION

The 2022 application cycle, saw a record number (643) of otolaryngology applicants vying for 361 residency positions. Within this competitive environment, programs received a record number of applications (427 on average) and allopathic applicants applied to a record number of programs: 84 on average.^1^

In response to rising application numbers and limitations imposed by the COVID pandemic on medical student away rotations, Otolaryngology Program Director Organization (OPDO) implemented an interview preference signaling system for both the 2021 and 2022 application cycles.^2,3^ Preference signaling is a mechanism that allows applicants to credibly signal their interests in an otherwise congested market, with the aim of facilitating mutually suitable matches between employer and applicant during the interview offer phase of the application process.^4,5^ While preference signaling has been utilized in the economics job market for prospective graduate students since 2006, the use in graduate medical education was pioneered by OPDO in the 2021 application cycle. Over the past two years, adoption of preference signaling has rapidly expanded within the GME community. We present the first year-over-year data comparison for signaling in this marketplace. As this concept remains relatively new to the medical education community, continued data sharing is required to enable both programs and prospective residents to understand its utility.

## METHODS

For the 2022 application cycle, applicants identified 4 programs they wished to send a signal. For the 2021 application cycle, 5 signals were provided. During both cycles, applicants were counselled not to send signals to visiting sub-internship and home programs. Likewise, these programs were informed not to expect signals from their home applicants or those that rotated with them. Additionally, applicants were asked to submit the name of another program that they would have signaled had they been given an additional signal to send. This additional program, designated the “comparator non-signaled” program, was used for quality evaluation purposes only. The comparator non-signaled program is assumed to resemble the characteristics of the officially signaled programs most closely.

Submissions were collected using a web-based interface at www.entsignaling.org. Each year, the signals were distributed to participating programs the same day Electronic Residency Application System (ERAS) released applications for viewing by programs.

The distribution of signals among programs was evaluated descriptively and in relationship to Doximity rankings.^6^ Mean number of signals were calculated for programs in ranking groups of 10. For programs that received separate signaling for their research track versus their non-research track, Doximity analysis was conducted using only signals from the non-research track to avoid potential for duplicate signaling by applicants to that program. Military programs were not included in the signaling initiative as their applicant population and their matching system has limited utility for preference signals. Military programs were thus removed from the calculation of the mean.

To assess the signaling process, a survey was sent the week before Match to applicants who participated in signaling. Along with inquiring about general attitudes towards the signaling process, applicants were asked the number of interview invitations received, how many invitations were garnered from signaled programs, non-signaled programs, and from the comparator non-signaled program.

Surveys were sent to otolaryngology training programs (program directors and program coordinators) after the Match to evaluate the impact of signaling on final Match results. Programs reported the number of matched applicants that came from their home medical school, completed a visiting in-person rotation within the current Match cycle, or sent a signal. Program directors were queried how they utilized the signals. Duplicate program survey responses were managed by eliminating responses that did not provide numerically correct data or provided responses that were not internally congruent. Students and program directors were also asked whether the signaling process should continue, and they were given space for unstructured comments. Respondents were not forced to answer all questions.

COVID pandemic circumstances made the two application cycles slightly different. However, in both the 2021 and 2022 application cycles, applicants had six “special” programs for analysis (Table 1). In 2021, applicants with home programs were afforded 5 signals plus their home program for analysis. Applicants without home programs received 5 signals plus a possible visiting sub-internship (visiting sub-internships were permitted only to those without home programs due to the COVID pandemic). In 2022, allowance for visiting sub-internship increased incrementally by one. Applicants were afforded 4 signals plus home plus one visiting sub-internship (or two visiting sub-internships permitted for applicants without a home otolaryngology program) for analysis. As this data was collected for quality assurance/ quality improvement purposes, the post hoc analysis of de-identified survey data was categorized as not human subjects research by the University of California, San Francisco Institutional Review Board and thus exempt from review.

**Table 1.** “Six Program” Preference Signaling Design

| Application cycle | 2021 |  | 2022 |  |
| --- | --- | --- | --- | --- |
| Applicant type | A | B | A | B |
| # Preference |  |  |  |  |
| signals | 5 | 5 | 4 | 4 |
| Home program | 1 | 0 | 1 | 0 |
| Visiting Sub-I | 0 | 1 | 1 | 2 |
| Total “signals” | 6 | 6 | 6 | 6 |

## RESULTS

### Signaling Data

In the 2021 and 2022 application cycle, there were 558 and 559 unique applicants respectively who signaled. There were 117 and 120 programs that participated in Match in 2021 and 2022 respectively (programs with more than one residency track were counted only once). Programs received a range of signals from 0-66, with 50% of signals going to 24% of programs (Figure 1), which was similarly found in 2021 (not shown). Programs of higher Doximity rank tended to receive more signals per program (Figure 2).

**Figure 1.**
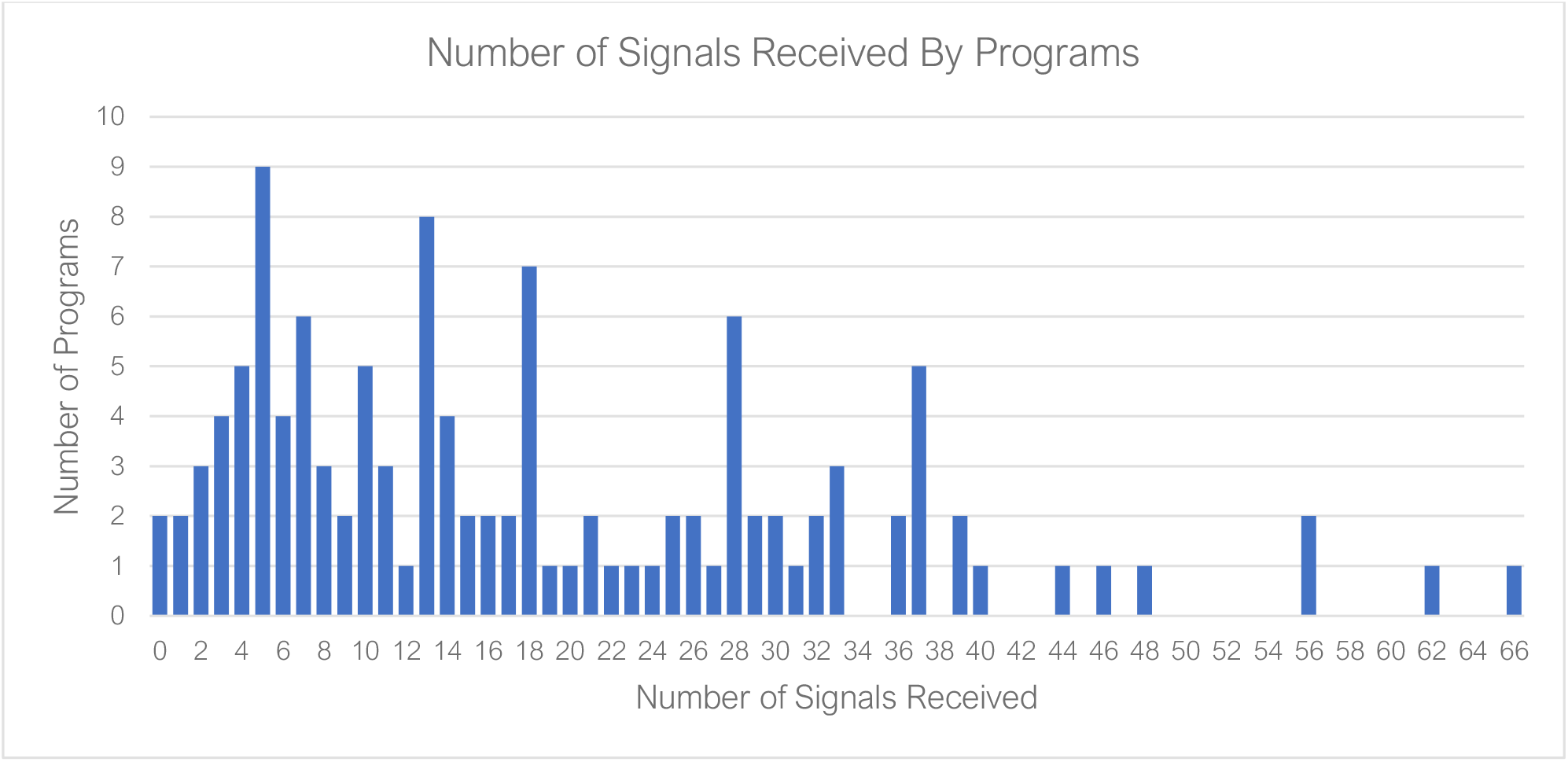
Histogram of number of progams and the number of singals received, 2022.

**Figure 2.**
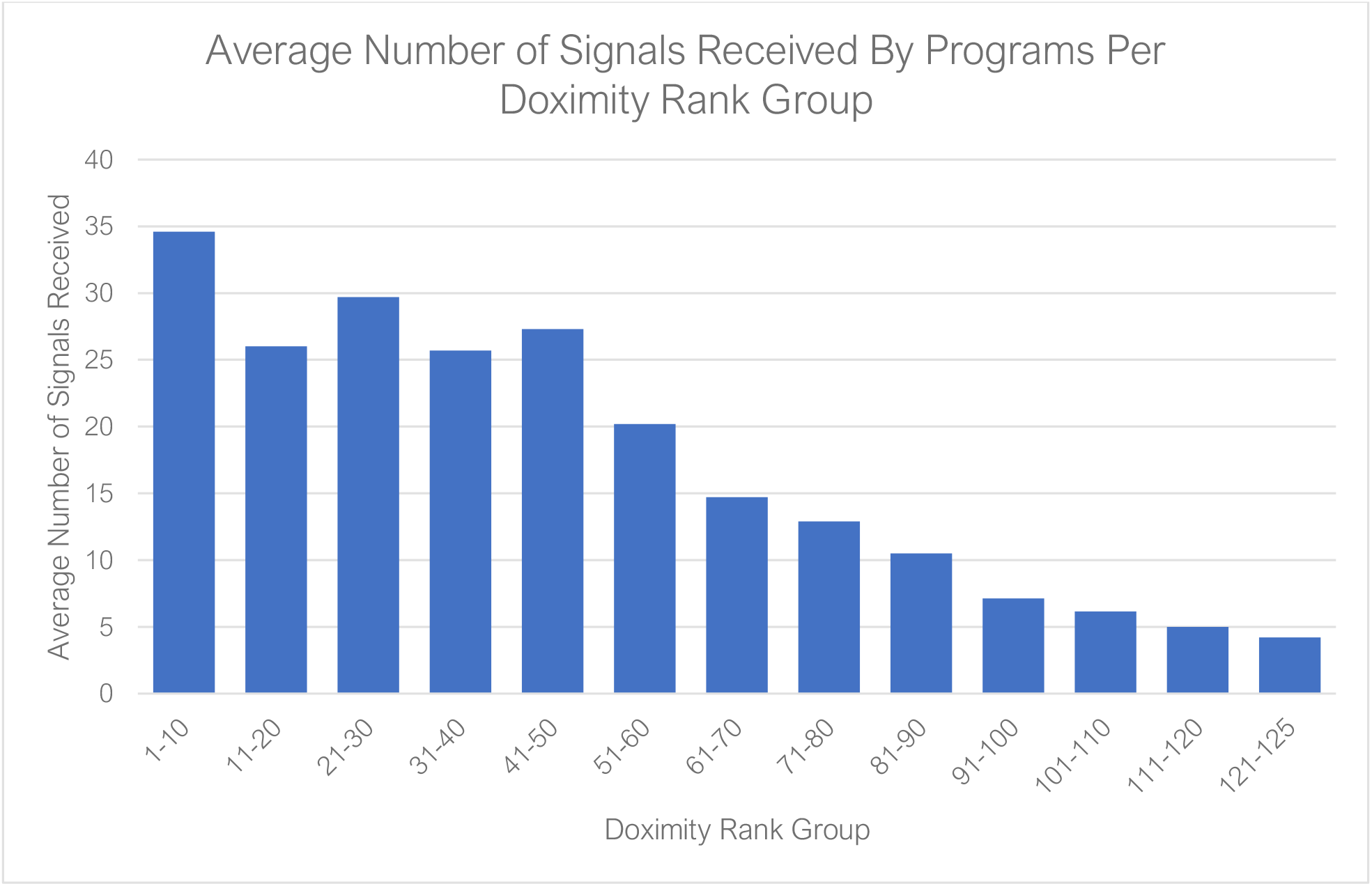
Average number of signals received by programs per Doximity rank group, 2022. Programs are grouped in bins of 10 to preserve anonymity.

### Applicant Survey Results

Survey response rate for applicants who participated in signaling in 2021 and 2022 was 42% (233/558) and 47% (263/559) respectively.

#### Interviews received

In 2022, among surveyed applicants, median and mean number of interview offers received was 11 and 12.8 respectively, with the spread between the 25^th^ percentile and 75^th^ percentile ranging from 7 to 18 invitations. Overall, 87% of surveyed applicants received an interview offer from at least one program they signaled (Figure 3). This compares to 93% in 2021, a year where signaling may have helped mitigate the severe restriction of visiting sub-internships. Applicants who received more interview offers from signaled programs also received more interview offers overall (Figure 4).

**Figure 3.**
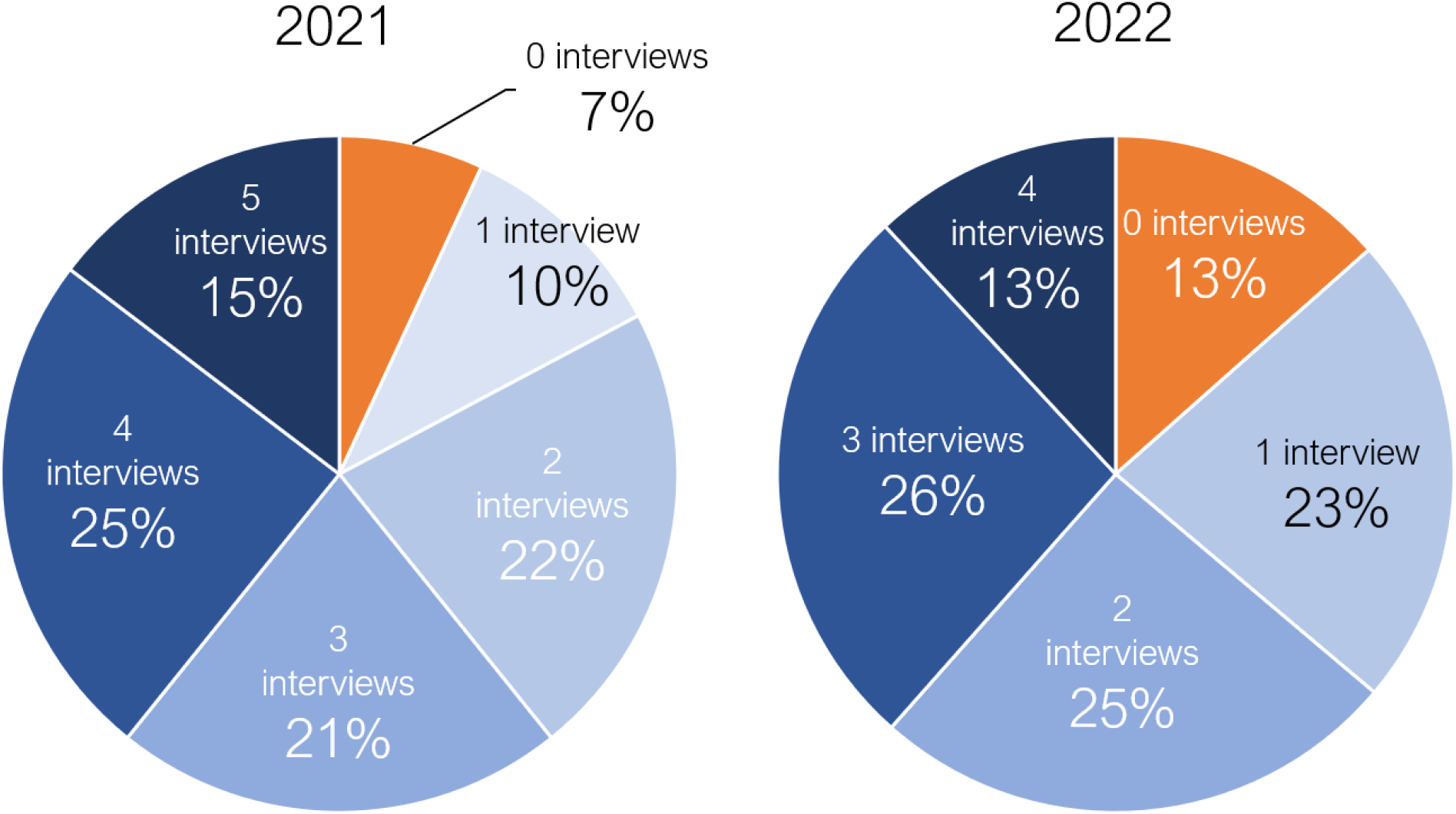
Pie chart of applicant-reported number of interviews received from signaled programs

**Figure 4.**
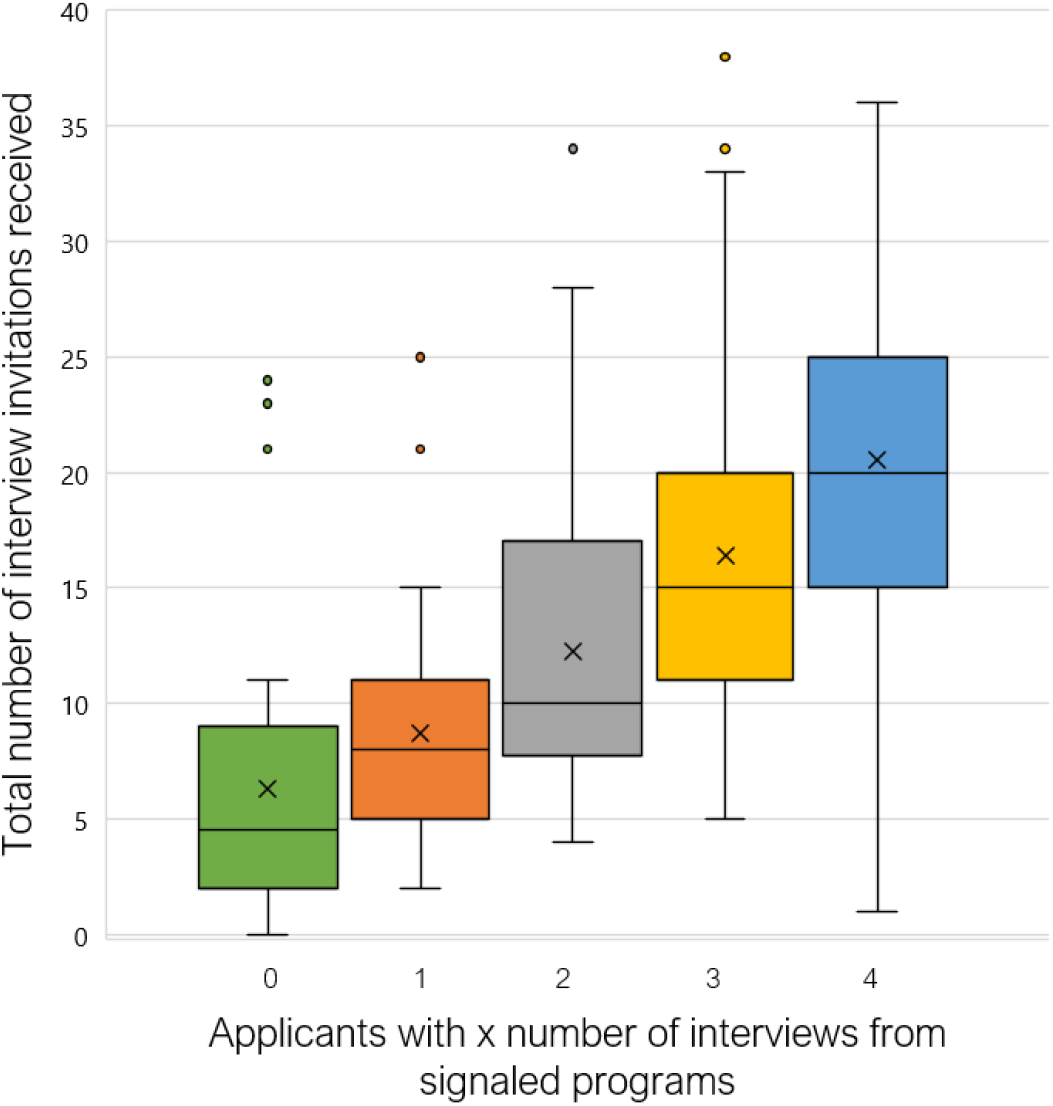
Total number of interviews received, 2022. Data is stratified by applicants who received 0, 1, 2, 3, or 4 interviews from signaled programs. Box plots represent 25-75%ile with horizontal line denoting median, X denotes mean, whiskers represent 95%tile.

#### Impact of signaling

The rate of receiving an interview offer was significantly higher for signaled programs (rate 50%) compared with both non-signaled programs (rate 13%; OR 6.7, 95% CI 5.9-7.7) and the comparator non-signaled program (rate 19%, OR 4.3, 95% CI 3.1-6.0; see Figure 5). While the overall offer rate was lower in Match 2022 than Match 2021, the multiplier effect of signaling remained similar: In both 2021 and 2022, applicants were 2.6 times more likely to get an interview from a signaled program than from the comparator non-signaled program. During the 2022 recruitment cycle, the survey also inquired specifically about interview rates among visiting sub-internship programs and home programs. Predictably, both options provided applicants with a high interview offer rate.

**Figure 5.**
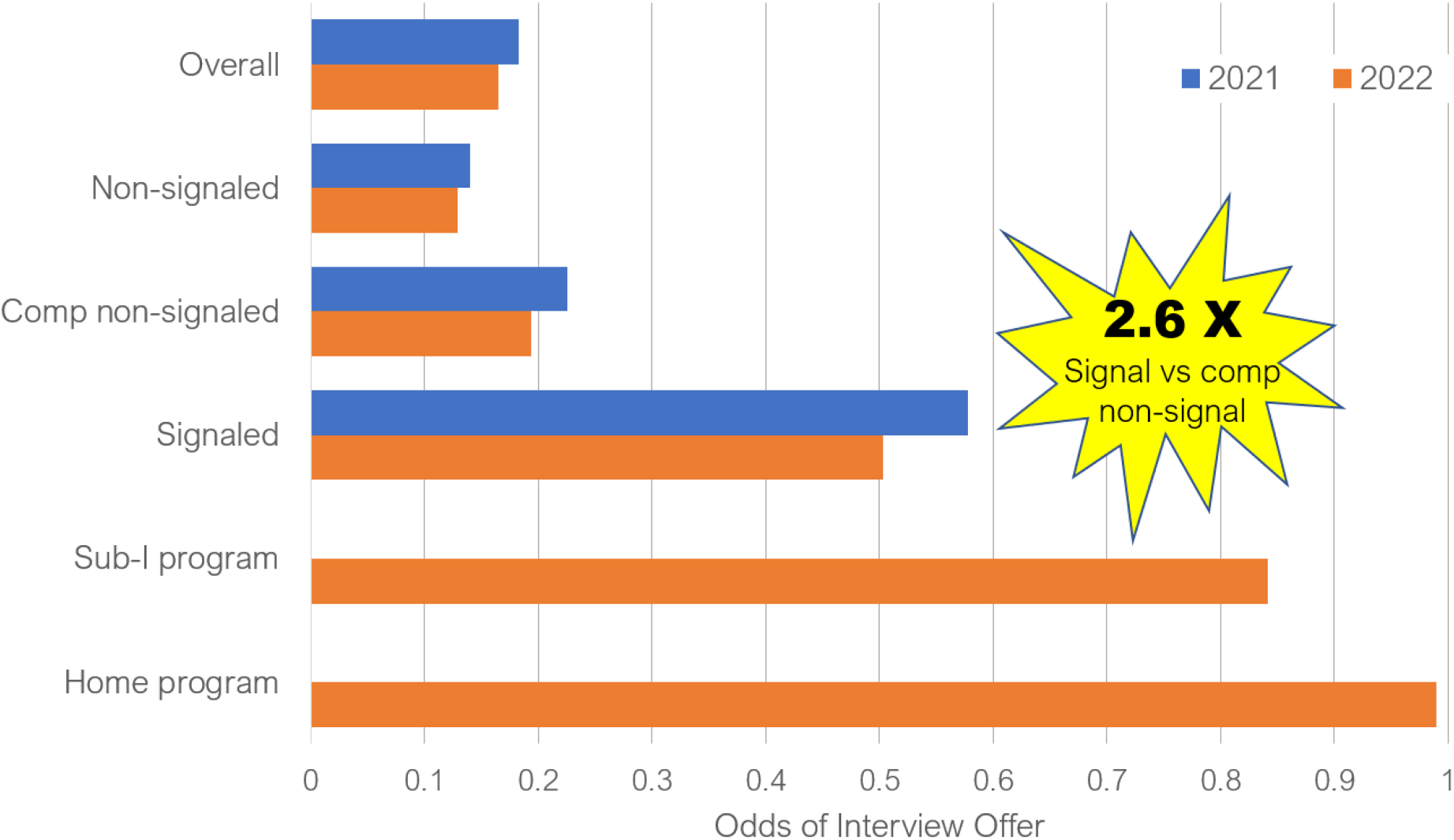
Interview offer rates, 2022. Rates are shown grouped by program attribute. Sub-I = sub-internship

To assess the impact of signaling across the spectrum of applicant competitiveness, we divided applicants into quartiles based on their overall likelihood of receiving an interview offer (# interview offers / #applications). Signals had a significant impact (P < .001) on the interview offer rate across all quartiles (Figure 6). A larger multiplier effect was seen among lower quartile applicants, suggesting that signaling had a greater positive impact on interview offer rate for the less competitive applicants. These findings are similar to last year’s results with the exception of the 4^th^ quartile where the multiplier effect was markedly increased in 2022. This is likely due to an overall decrease in interview offer rate from non-signaled programs within this quartile.

**Figure 6.**
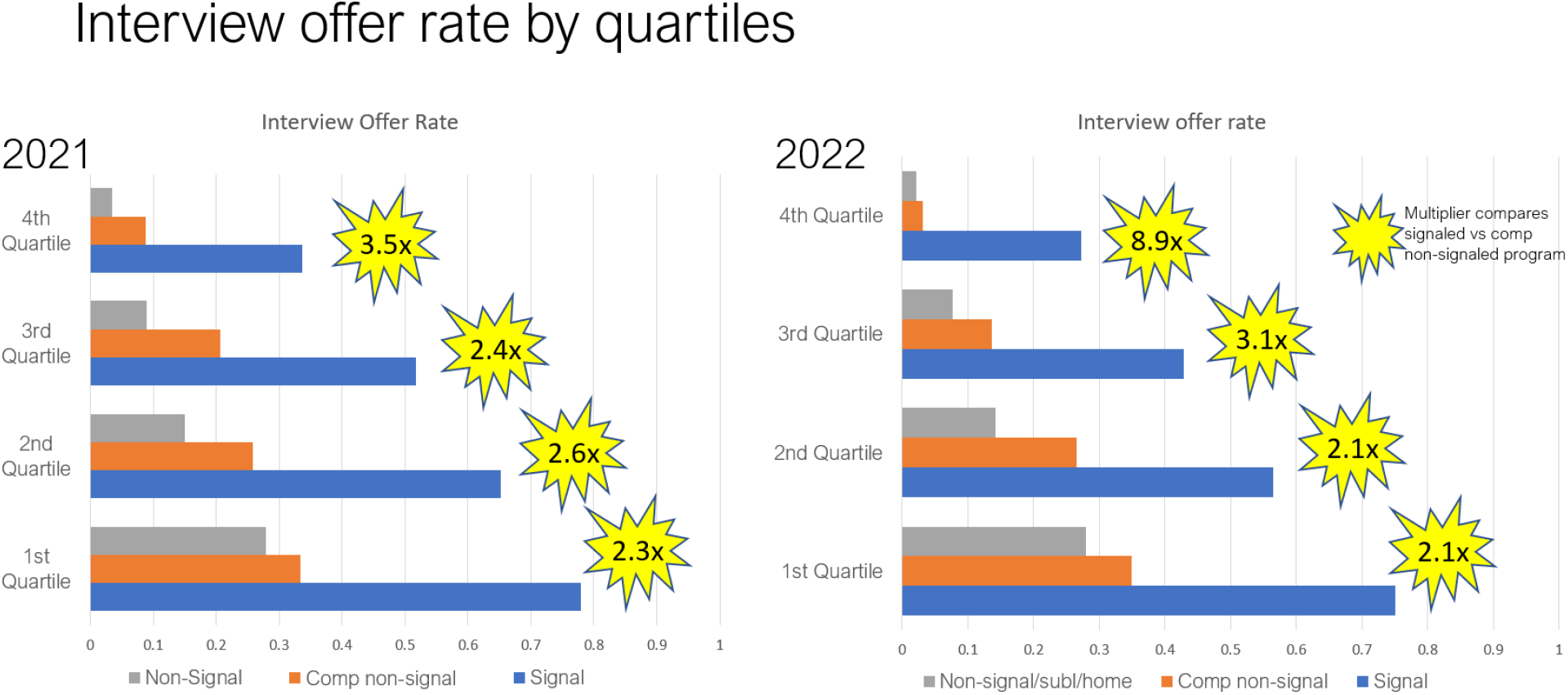
Interview offer rate by quartiles, 2021 and 2022. Comp non-signal = comparator non-signaled program.

### Program survey results

Survey response rate for programs was 52% (62/117) and 53% (64/120) for 2021 and 2022 respectively.

#### Use of signals in residency application review

In 2022, program directors were queried as to how the signals were utilized in the applicant recruitment process. Programs were permitted to provide multiple answers (Figure 7). Most programs incorporated signals into the initial review or used signals as a tie breaker for interview invitations. Four programs responded they did not use signaling (two cited a communication error and two programs merely cited they did not use the signals they were provided). Five programs incorporated signaling information into the development of their rank lists.

**Figure 7.**
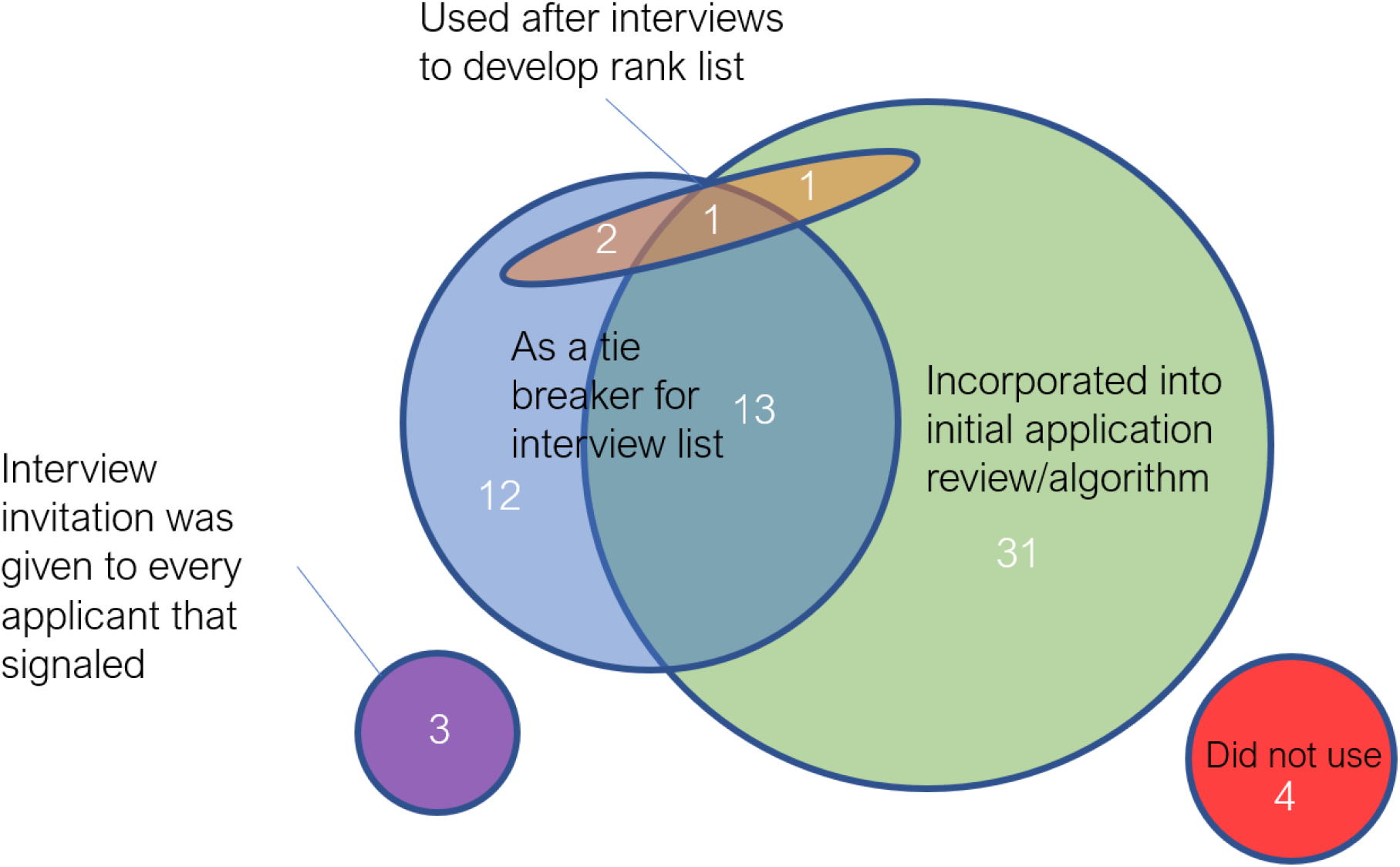
Venn diagram of how programs used signals received, 2022.

#### Positions filled by signals

Programs reported that 60% and 70% of their matches in 2021 and 2022 respectively were from applicants that either signaled, completed a visiting sub-internship, or were from their own program (Figure 8).

**Figure 8.**
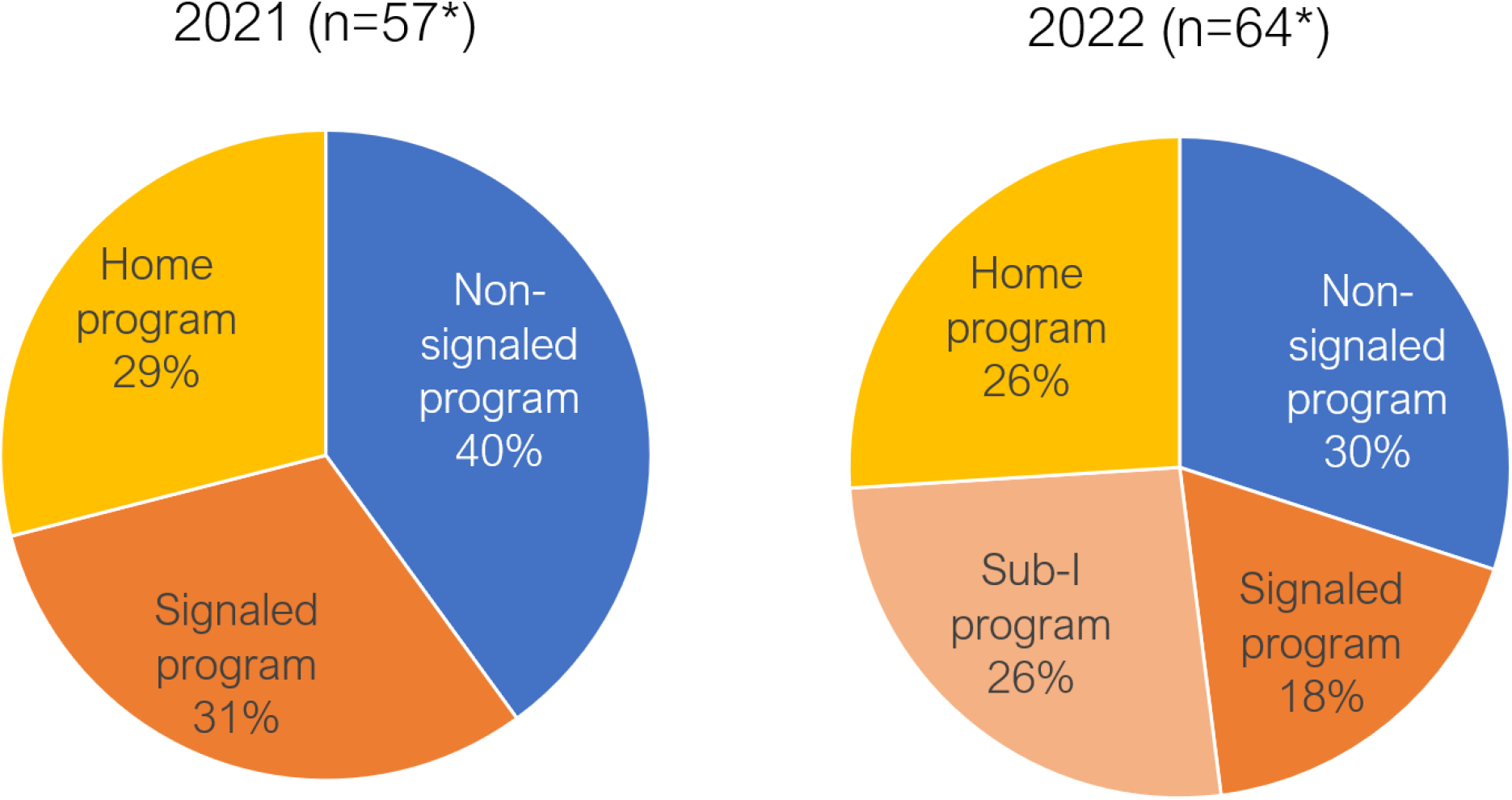
Signaling characteristics of applicants who filled positions, as reported by programs. *entries from programs excluded if data were omitted or if reported match data were discrepant. Sub-I = sub-internship

### Applicant and Program Opinions on Preference Signaling

Signaling was viewed favorably by most surveyed applicants and programs. Nearly 90% of both constituents wished to continue with the signaling initiative. More than 60% of applicants and more than 70% of programs agreed or strongly agreed with being satisfied with signaling (Figure 9).

**Figure 9.**
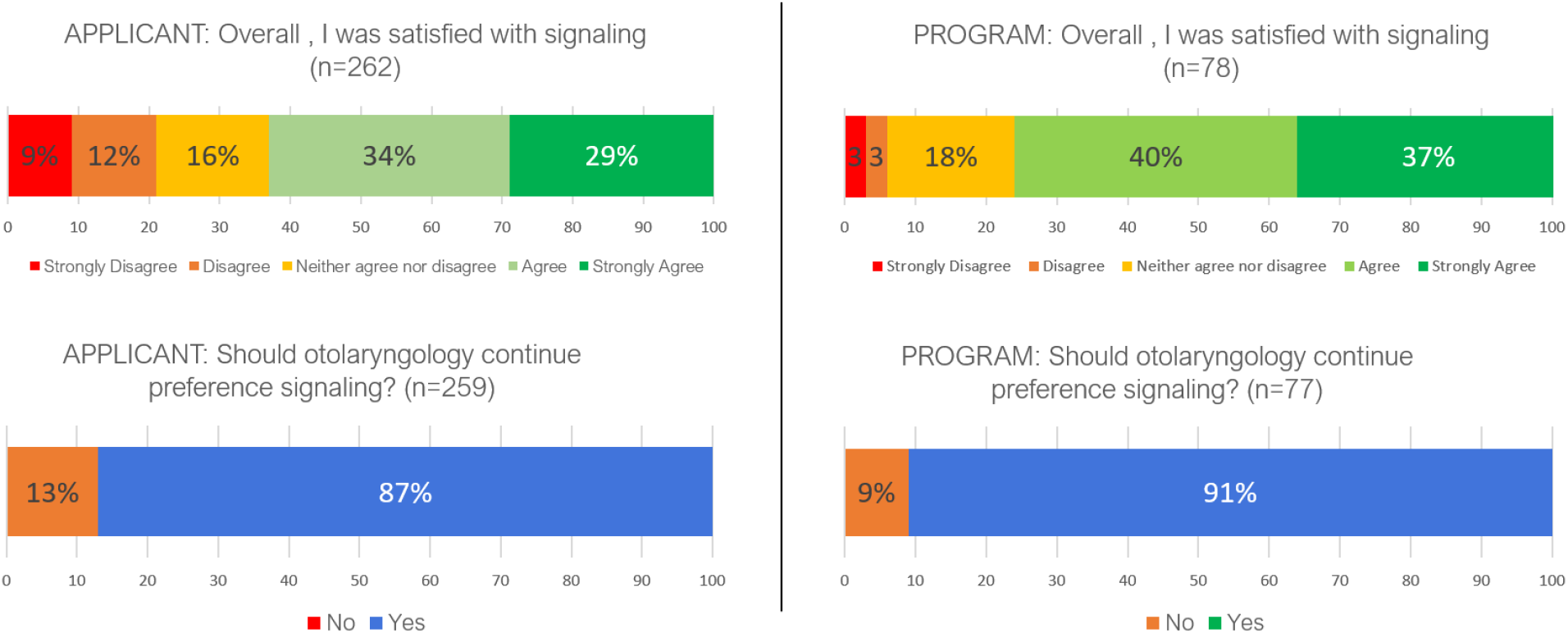
Applicant and program opinions of preference signaling, 2022.

## DISCUSSION

Preference signaling is a mechanism intended to allow applicants the ability to demonstrate credible interest in receiving an interview from selected programs. With the large rise of applications submitted by each applicant, an individual application to a program is diluted by the large number of applications each program receives. In the 2021 application cycle, otolaryngology was the first specialty in graduate medical education to implement a preference signaling system for residency interviews.^1^

Survey results from this initial experience demonstrated that signals markedly increased the likelihood of receiving an interview offer. The impact of signals was greatest for applicants who struggled most to obtain interview offers. This finding continued to hold true for the 2022 application cycle.

For the applicant, receiving an interview from a specific program is virtually assured if such program is the applicant’s home program or if the applicant completed a visiting sub-internship at that program. This advantages applicants from medical schools with otolaryngology programs as well as those applicants able to secure coveted visiting sub-internship rotations. This translates to match outcomes that strongly favor applicant-program pairings with home program or visiting sub-internship relationships. Such biases may perpetuate equity challenges inherent in medical school matriculation as well as the process involved in obtaining visiting otolaryngology sub-internships. Preference signaling helps applicants garner interviews with programs in the absence of any pre-existing relationship, with a 260% interview invitation rate boost. This impact is robust and present across the demographic categories of gender and self-identified URIM status (pre-published data).

With this early success, preference signaling was incorporated into the 2022 Match through a collaboration with the Association of American Medical Colleges (AAMC) and ERAS pilot program involving internal medicine, general surgery, and dermatology. Urology also adopted preference signaling, independent of AAMC and ERAS. After favorable initial impressions, this pilot has been expanded to include 15 specialties for the 2023 application cycle.^7^

Signaling may provide a pathway for additional modifications to the residency application process. One solution proposed to address the progressive increase in the number of applications submitted per applicant is to implement an application cap. Models to evaluate the impact of a cap are limited by the ability to predict applicant behavior. However, applicant strategies used to choose programs to signal are similar to strategies that would be employed with an application cap. Evaluation of match positions filled with a limited “6-program” system models a very low application cap. The finding that >70% of programs filled in this scenario suggests that in a specialty like otolaryngology, an application cap of 5 times the number would very likely result in matching of all residency slots and reduce the surplus of 40+ applications per applicant. Furthermore, programs could devote more time to apply the principles of holistic review with a decreased application load. In the 2023 application cycle, orthopedic surgery and obstetrics-gynecology are piloting higher number of preference signals (30 and 18 respectively)^6^ that, if all programs fill with signaled and home applicants, may validate an application cap approach.

Even in the absence of an application cap, given the appropriate number of signals, candidates could feel less compelled to over-apply. More competitive candidates would realize cost savings, confident that their applications would be suitably noticed with signaling. Less competitive applicants could continue to pursue an unrestrained application strategy if they so wish. Thus far, a reduction in application submissions has not been realized, however this was not the original intent of the preference signaling process. If downward pressure on over application is desired, transparency of signaling data outcomes is required to feedback to applicants the effects of signaling. Upward adjustment of the number signals provided to each applicant may be needed to provide the reasonably competitive applicants the statistical and psychological reassurance of match success.

Continued messaging regarding the intent of the signaling process is needed to combat misperceptions. Preference signaling is intended to be used to facilitate selection of candidates to interview. A signal is not intended to guarantee an interview. Programs will use signals variably depending upon their recruitment strategy. This is not unlike the variable use of any application metric. Match rank lists created by both the applicant and the program should match their professional and departmental goals respectively to achieve optimal outcomes, regardless of interview preference signaling. During the first 2 years of preference signaling, applicants were instructed not to signal their home program or visiting sub-internship program to avoid coercion of a forced “loyalty pledge.” Signaling was an appropriate equity strategy in years where opportunities for visiting sub-internships were constrained. However, if in-person visiting sub-internship scheduling returns to pre-pandemic levels, this guidance may need modification.

Limitations of this study include an incomplete survey response rate and potential for biased sample of respondents. Generalizability of otolaryngology preference signaling results to other specialties may vary due to differences in the size of other programs, number of signals provided to each applicant, applicant characteristics, and differing recruitment climate.

## CONCLUSIONS

Preference signaling for otolaryngology residency interviews demonstrates a promising mechanism for applicants to better influence visibility to programs during the application cycle. This impact is consistent over 2 application cycles. Continued iterations may help reduce the application burden for applicants.

## Data Availability

All data produced in the present study are available upon reasonable request to the authors

## Acknowledgments

The authors thank Emily Maurer, senior administrator at the American College of Surgeons, for her invaluable contributions to the execution of the otolaryngology signaling program. They also thank the Otolaryngology Program Directors Organization and the councils of the Society of University Otolaryngologists and the Association of Academic Departments of Otolaryngology for their support of the signaling program.

